# Mapping access to basic hygiene services in low- and middle-income countries: A cross-sectional case study of geospatial disparities

**DOI:** 10.1101/2020.08.07.20169995

**Authors:** Weiyu Yu, Robert E. S. Bain, Jie Yu, Victor Alegana, Winfred Dotse-Gborgbortsi, Yi Lin, Jim A. Wright

**Affiliations:** School of Geography and Environmental Science, University of Southampton, Building 44, Highfield, Southampton S017 1BJ, UK; Division of Data, Analysis, Planning and Monitoring, United Nations Children’s Fund (UNICEF), 3 United Nations Plaza, New York, NY 10017, United States; College of Environmental Science and Engineering, Tongji University, Shanghai 200092, China; Population Health Unit, Kenya Medical Research Institute (KEMRI) - Wellcome Trust Research Programme, Nairobi, Kenya; College of Surveying and Geo-Informatics, Tongji University, Shanghai 200092, China

**Keywords:** Basic hygiene, Handwashing, water and soap, WASH, ensemble model, machine learning

## Abstract

Handwashing with water and soap, is among the most a cost-effective interventions to improve public health. Yet billions of people globally lacking handwashing facilities with water and soap on premises, with gaps particularly found in low- and middle-income countries. Targeted efforts to expand access to basic hygiene services require data at geospatially explicit scales. Drawing on country-specific cross-sectional Demographic and Health Surveys with georeferenced hygiene data, we developed an ensemble model to predict the prevalence of basic hygiene facilities in Malawi, Nepal, Nigeria, Pakistan and Uganda. The ensemble model was based on a multiple-level stacking structure, where five predictive modelling algorithms were used to produce sub-models, and a random forest model was used to generalise the final predictions. An inverse distance weighted interpolation was incorporated in the random forest model to account for spatial autocorrelation. Local coverage and a local dissimilarity index were calculated to examine the geographic disparities in access. Our methodology produced robust outputs, as evidenced by performance evaluations (all R^2^ were above 0.8 with the exception of Malawi where R^2^ = 0.6). Among the five study countries, Pakistan had the highest overall coverage, whilst Malawi had the poorest coverage. Apparent disparities in basic hygiene services were found across geographic locations and between urban and rural settings. Nigeria had the highest level of inequalities in basic hygiene services, whilst Malawi showed the least segregation between populations with and without basic hygiene services. Both educational attainment and wealth were important predictors of the geospatial distribution of basic hygiene services. By producing geospatially explicit estimates of the prevalence of handwashing facilities with water and soap, this study provides a means of identifying geographical disparities in basic hygiene services. The method and outputs can be useful tools to identify areas of low coverage and to support efficient and precise targeting of efforts to scale up access to handwashing facilities and shift social and cultural norms on handwashing.

## 1. Introduction

Hand hygiene is a measure of personal hygiene and a cost-effective non-pharmaceutical intervention to improve public health by preventing the transmission of infectious diseases (Warren-gash *et al*. 2012, Loughnan *et al*. 2015, White *et al*. 2020). Handwashing with water and soap has been found particularly effective in reducing the spread of influenza (Talaat *et al*. 2011), respiratory tract viruses (Roberts *et al*. 2000, Rabie and Curtis 2006, Jefferson *et al*. 2011) and diarrhoeal diseases (Fewtrell *et al*. 2005, Huang and Zhou 2007, Ejemot *et al*. 2008, Wolf *et al*. 2018, Dey *et al*. 2019), as it is likely to interrupt transmission via fomites and to a certain extent close contact routes such as droplets (Warren-gash *et al*. 2012). In light of disease and epidemic persistence throughout history, practicing of good hand hygiene is often recommended in public health guidelines and has remained a key component of personal level protection strategy during the recent pandemic events (World Health Organization 2009, 2020, WHO and UNICEF 2020). The World Health Organization (WHO)/United Nations Children’s Fund (UNICEF) Joint Monitoring Programme for Water Supply, Sanitation and Hygiene (JMP) identified handwashing with water and soap, referred to as a basic hygiene service, as one of the top priorities for monitoring of progress towards the Sustainable Development Goals (SDG) Target 1.4 and Target 6.2 (WHO and UNICEF 2018).

Despite its importance, significant efforts are still required to increase the prevalence of hand hygiene with water and soap in many low- and middle-income countries (LMICs), particularly in poor and marginalised settings where people are disadvantaged by a lack of basic infrastructure and education (Loughnan *et al*. 2015, Renzaho 2020). In a recent study, Brauer et al. (2020) estimated approximately two billion people globally still lacked access to basic handwashing facilities with water and soap at home in 2019, with barriers to universal access mostly in LMICs. At subnational level, large disparities in access were found across geographic locations and between urban and rural settings (Brauer *et al*. 2020, Jiwani and Antiporta 2020). Access to a hygiene facility with water and soap can be extremely low even in urban areas in countries such as Malawi where local cleansing agents (e.g. ash, mud, etc.) are often chosen over soap as cheaper and more acceptable alternatives (Nguyen 2015), although their effects on preventing disease transmission remain uncertain (Paludan-Müller *et al*. 2020). In this context, implementing hand hygiene interventions in response to an emergency situation such as the ongoing coronavirus diseases 2019 (COVID-19) pandemic can be challenging (Jiwani and Antiporta 2020). Such circumstances call for rapid resource deployments by governments and development partners to scale up access to hand hygiene facilities with water and soap and shift social and cultural norms on handwashing (UNICEF and WHO 2020), which in turn requires knowledge about hand hygiene facilities and behaviours at the sub-provincial level.

Nationally representative household surveys such as the Demographic and Health Surveys (DHS) and the Multiple Indicator Cluster Surveys (MICS) are often the key sources of data on hand hygiene for low- and middle-income settings. Being designed for multiple purposes, DHS and MICS surveys rely on rapid observations of hygiene facilities to balance cost-effectiveness and representativeness (Ram 2013). In comparison with other small-scale data collected via rigorous methods such as structured observations on behaviours, these surveys cover larger geographic extents and a wider range of demographic and health-related characteristics. They are therefore widely used as proxy indicators for actual behaviour (Ram 2013, Loughnan *et al*. 2015). However, whilst household surveys are typically representative at subnational level, for example, at provincial (the first administrative) level and between urban and rural areas, more geographically disaggregated estimates are often lacking due to survey sampling design and confidentiality protection (ICF International 2012). Other recent efforts to estimate coverage of basic hygiene services have also been limited to the national or the first subnational administrative levels (Brauer *et al*. 2020). To gain a more detailed view of access to basic hygiene services from household surveys and in order to inform development policy, resource deployment and intervention implementation, local coverage estimates should be produced for all locations within a country.

One approach to producing geospatially explicit estimates for all locations is through Bayesian geostatistical modelling (Diggle *et al*. 1998), given its long history in mapping other demographic and public health indicators from household survey data (Gosoniu *et al*. 2010, Magalhães and Clements 2011, Osgood-Zimmerman *et al*. 2018, Reiner *et al*. 2018, Dwyer-Lindgren *et al*. 2019, Mayala *et al*. 2019, Mosser *et al*. 2019). The Bayesian geostatistical model predicts the prevalence of a target indicator at unobserved locations by quantifying the relationship between the prevalence at observed locations with potential predictive covariates, whilst accounting for spatial dependence via a covariance matrix of a Gaussian process with location-specific random effects (Karagiannis-Voules *et al*. 2013, Lai *et al*. 2013). A common approach to implementing such Bayesian geostatistical models is through the Markov Chain Monte Carlo (MCMC) algorithm in computational software such as WinBUGS (Medical Research Council Biostatistics Unit, Cambridge & Imperial College London, London, UK). Because of the large covariate matrices involved, its implementation often suffers drawbacks such as lack of convergence, high storage requirements, and high computational cost (Lai *et al*. 2013, Mayala *et al*. 2019). There have been efforts to address these issues, among which the Integrated Nested Laplace Approximation (INLA) methodology (Rue *et al*. 2009) has become increasingly popular in mapping demographic and health-related indicators from household survey data (Osgood-Zimmerman *et al*. 2018, Reiner *et al*. 2018, Dwyer-Lindgren *et al*. 2019, Mayala *et al*. 2019, Mosser *et al*. 2019). INLA constructs a triangulation (known as a mesh) over the study area, and computes the spatial autocorrelation structure of the dataset at the mesh vertices using a stochastic partial differential equation (SPDE) approach (Lindgren *et al*. 2011). In comparison with MCMC methods, INLA provides a significant speed boost with high accuracy, and has been implemented in R environment through the ‘R-INLA’ package (Lindgren and Rue 2015). However, being computationally efficient, such an approach generates an approximation with potential drawbacks, such as the boundary effect in the covariance approximation due to boundary conditions of the SPDE (Lindgren *et al*. 2011). As an alternative to geostatistical models, machine learning predictive models are widely employed in geospatial mapping applications (Wang *et al*. 2010, Massada *et al*. 2012, Stevens *et al*. 2015, Pearce *et al*. 2016, Naghibi *et al*. 2017, Yu *et al*. 2019). In comparison with geostatistical models, machine learning predictive models often require fewer statistical assumptions and can be flexibly automated (Hengl *et al*. 2018). Whilst they suffer the major drawback of ignoring spatial autocorrelation, a recent study (Hengl *et al*. 2018) has incorporated geographical proximity effects into a Random Forest (RF) (Breiman 2001) model by calculating buffer distances from sample observation points. This method, known as random forest for spatial predictions (RFsp), has generated comparable results to geostatistical models, but may be impractical for large data applications, given the intensive computational costs associated with buffer distances for all sample observation points.

In this study, we adopted an ensemble model of machine learning algorithms to produce geospatially explicit estimates for all locations within the case study countries. Drawing on the rationale behind RFsp, we incorporated geographical proximity effects into the final model prediction to account for spatial autocorrelation. The main objectives of this study are (1) to produce geospatially explicit estimates of basic hygiene prevalence across the case study countries; (2) to examine the applicability of the ensemble machine learning model for such applications; (3) to examine the relative importance of covariates for predicting basic hygiene services; and (4) to quantify geographic disparities in access to basic hygiene services using the resultant geospatial estimates of basic hygiene services.

## 2. Materials and Methods

### 2.1 Study countries and sample data

In this study, we selected five case study countries for which the most recent (post-2015) georeferenced DHS datasets and recent geospatial datasets characterising factors affecting access to basic hygiene services, particularly poverty were available. Selected countries were classified as either low- or middle-income by the World Bank (World Bank 2020a), and comprised Malawi, Nigeria and Uganda in sub-Saharan Africa, and Nepal and Pakistan in South Asia. Estimated national coverage of basic hygiene services in these five countries ranges from 9% in Malawi to 63% in Pakistan (Brauer *et al*. 2020). **Figure 1** shows the locations of the five study countries and clusters (i.e. the groupings of households participated in the DHS campaign) from the latest DHS.

**Figure 1.**
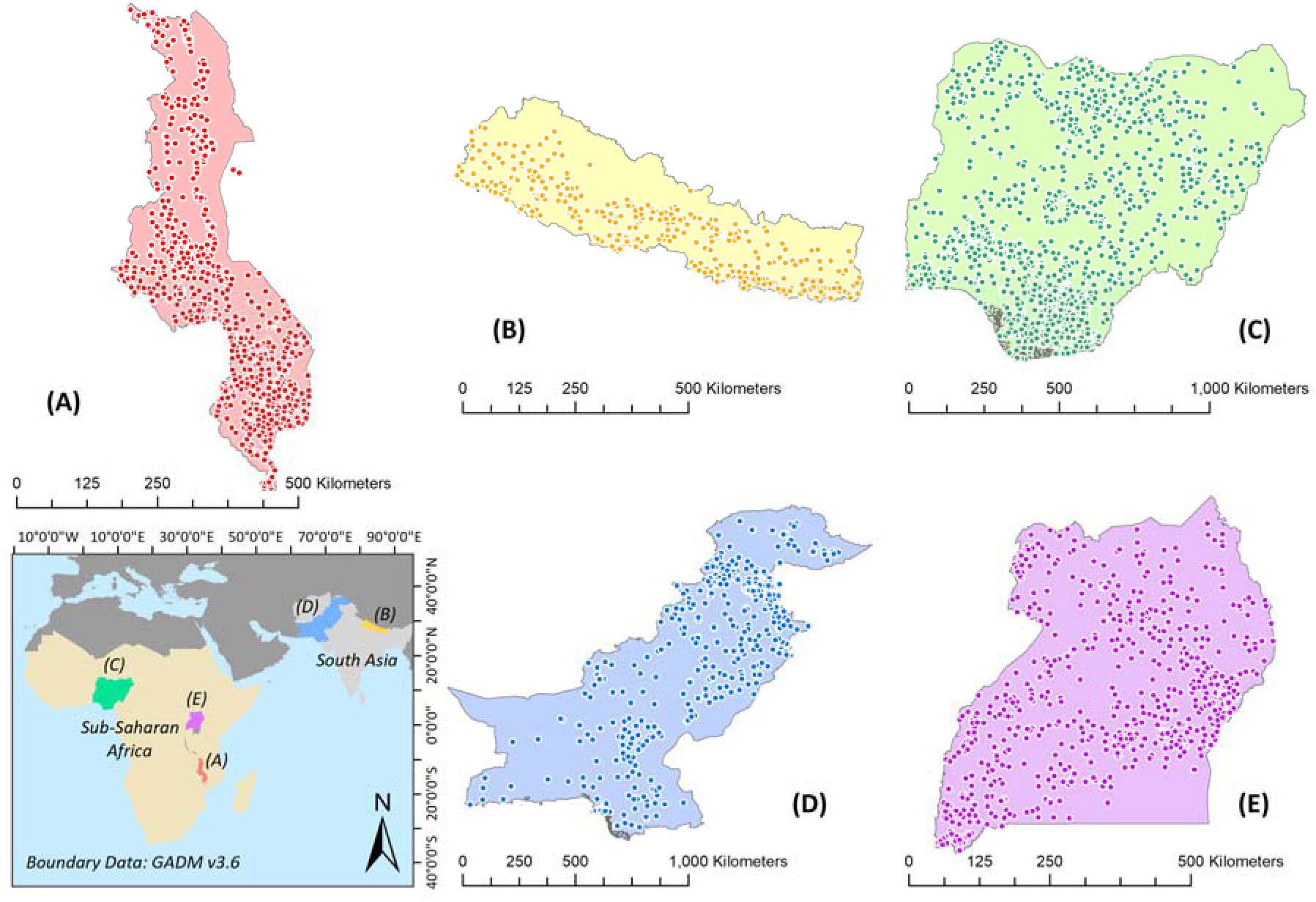
Map showing the geographic locations of (A) Malawi, (B) Nepal, (C) Nigeria, (D) Pakistan and (E) Uganda, together with cluster point locations (coloured dots) for the latest DHS.

We obtained country-specific standard DHS data from the most recent round (Phase VII) via the DHS Program portal (https://www.dhsprogram.com/data/available-datasets.cfm). covering household survey and geographic data for Malawi DHS 2015-16, Nepal DHS 2016, Nigeria DHS 2018, Pakistan DHS 2017-18 and Uganda DHS 2016. These DHS household surveys included observation-based measurements of handwashing facilities. During the survey, interviewers asked respondents to show them the fixed place or mobile station used most often by household members for handwashing. Where feasible, presence of water and cleansing agent were directly observed at the place for handwashing and recorded by the interviewer. The derived data therefore allow us to capture the proportion of population living in a household with an observed fixed place or mobile station for handwashing where both water and soap are available.

The DHS household surveys were based on a stratified two-stage cluster design (Croft *et al*. 2018) and weights are used to adjust for non-response and disproportionate sampling in regions with smaller and larger populations. In this study, the DHS household weight was multiplied by the *de jure* number of household members in order to produce estimate for the proportion of the population. Derived information on individuals living in a household with access to basic hygiene was aggregated to cluster level and then combined with the DHS geographic data. Generated tabular data therefore included the outcome variable - the proportion of *de jure* population living in a household with an observed handwashing facility where water and soap were present - alongside Global Positioning System (GPS) coordinates (longitude and latitude) for the DHS clusters. These GPS coordinates were provided with displacement of up to 2km for urban locations and up to 5km for rural locations (and up to 10km for every 100^th^ rural location) to protect respondent confidentiality (Burgert, Colston, *et al*. 2013, Perez-Heydrich *et al*. 2013). By overlaying these georeferenced cluster points with pre-processed geospatial covariate layers (detailed in the next subsection), those with implausible coordinates, located within the same 5km-grid cell, within an identifiable water body, or outside the boundary of study country were excluded. **Table 1** summarises the characteristics of the georeferenced DHS household survey data describing basic hygiene services and the number of cluster points included as the final sample for each study country.

**Table 1.** Characteristics of the obtained country-specific DHS data describing basic hygiene services

|  | Malawi | Nepal | Nigeria | Pakistan | Uganda |
| --- | --- | --- | --- | --- | --- |
| Sample size (number of households) | 26,361 | 11,040 | 40,427 | 14,540 | 19,588 |
| Number of households with hygiene information | 26,361<br>(100.0%) | 11,040<br>(100.0%) | 40,427<br>(100.0%) | 14,535<br>(>99.9%) | 19,588<br>(100.0%) |
| Number of households without hygiene information or missing data | 0<br>(0.0%) | 0<br>(0.0%) | 0<br>(0.0%) | 5<br>(<0.1%) | 0<br>(0.0%) |
| Number of households having water and soap at the observed place for handwashing | 2,596<br>(9.8%) | 4,991<br>(45.2%) | 11,042<br>(27.3%) | 8,892<br>(61.2%) | 4,788<br>(24.4%) |
| Survey respondent: |  |  |  |  |  |
| Household's head | 14,618<br>(55.5%) | 7,425<br>(67.3%) | 31,533<br>(78.0%) | 4,514<br>(31.1%) | 12,481<br>(63.7%) |
| Wife/husband of household's head | 9,898<br>(37.5%) | 2,331<br>(21.1%) | 6,342<br>(15.7%) | 6,344<br>(43.6%) | 5,690<br>(29.0%) |
| Son/daughter of household's head | 1,021<br>(3.9%) | 586<br>(5.3%) | 1,713<br>(4.2%) | 1,822<br>(12.5%) | 803<br>(4.1%) |
| Total number of household members in the sampled households | 119,326 | 47,026 | 186,450 | 98,895 | 89,202 |
| Total number of clusters | 850 | 383 | 1,389 | 561 | 696 |
| Number of clusters with geographic data | 850<br>(100.0%) | 383<br>(100.0%) | 1,359<br>(97.8%) | 560<br>(99.8%) | 685<br>(98.4%) |
| Number of clusters located within the spatial extent of the covariate layers (final sample) | 783<br>(92.1%) | 374<br>(97.7%) | 1,323<br>(95.2%) | 558<br>(99.5%) | 637<br>(91.5%) |

### 2.2 Geospatial covariates

We obtained geospatial datasets from publicly available data sources to create candidate covariate layers (as detailed in **Table S.1** in **Supplementary Material 1**). These covariates were selected for their potential to predict basic hygiene prevalence based on theory and existing literature on either factors influencing hygiene practices (Luby and Halder 2008, Wolf *et al*. 2019) or mapping of water, sanitation and hygiene (WASH) access (Gething *et al*. 2015, Mayala *et al*. 2019, Yu *et al*. 2019, Brauer *et al*. 2020, Ekumah *et al*. 2020).

For socio-economic factors, we included geospatial covariates characterising population density accessibility to urban centres, and proximity to land features (cultivated areas, major roads, and road intersections) which may reflect the socio-economic environment and in turn potentially affect access to hygiene facilities. In addition, since piped water has previously been applied as a covariate in mapping global access to basic hygiene (Brauer *et al*. 2020), and since the presence of handwashing items may relate to household amenities given the need for handwashing after toilet use for example (Wolf *et al*. 2019), we therefore included existing modelled map surfaces of improved water sources and open defecation (lack of sanitation) from the DHS Spatial Data Repository (Gething *et al*. 2015, Mayala *et al*. 2019) as proxies. We also included literacy as an educational attainment outcome, also previously included as a component of a socio-demographic index in predicting coverage of handwashing with water and soap (Brauer *et al*. 2020). Moreover, since wealth has previously been shown to correlate with handwashing (Luby and Halder 2008), we selected a series of indices measuring the spatial distribution of poverty for countries where available. This included the percentage of people living on less than $1.25 per day and $2 per day (Tatem *et al*. 2013), the percentage of people living in poverty defined by a Multidimensional Poverty Index (MPI) (Tatem *et al*. 2013, Alkire *et al*. 2015), the Wealth Index (WI) and the International Wealth Index (IWI) (Bosco *et al*. 2018). Furthermore, we calculated another global poverty index following the methodology described in Elvidge et al. (2009) using recent population and night-time lights datasets obtained from WorldPop (https://www.worldpop.org/) and the Earth Observation Group (EOG) at the National Oceanic and Atmospheric Administration (NOAA)/National Geophysical Data Center (NGDC) (https://www.ngdc.noaa.gov/eog/index.html). Floating point radiance values from the Visible Infrared Imaging Radiometer Suite (VIIR) Day/Night Band (DNB) sensor were rescaled to 1-100 before calculation so as to avoid any numeric difficulties. Since satellite-observed stable night-time lights have been found to correlate with economic activities (Pinkovskiy and Sala-i-Martin 2016), we also included stable night-time lights directly as a proxy locational metric of economic status. Further geospatial data characterising health conditions were also selected, including women with anaemia, child mortality (neonatal mortality and under-5 mortality) and stunting, underweight and wasting as objective undernutrition measures reflecting socio-economic status, infectious disease prevalence (diarrhoea, HIV, lower respiratory infection), and thereby poor hygiene (Fewtrell *et al*. 2005, Luby *et al*. 2005, Huang and Zhou 2007, Ejemot *et al*. 2008, Luby and Halder 2008, Curtis *et al*. 2011, Jefferson *et al*. 2011, Rah *et al*. 2015). For environmental factors, we included geospatial covariates describing elevation, slope, precipitation, aridity and potential evapotranspiration (PET) previously applied in predictive mapping of water sources and sanitation facilities (Gething *et al*. 2015, Mayala *et al*. 2019, Yu *et al*. 2019).

For all geospatial covariates, we selected data sources from as close to the present year as possible (mostly post-2015 and close to the survey year), with the exceptions of some environmental covariates which were either long-term means (e.g. precipitation, aridity, and potential evapotranspiration) or assumed to be temporally static (e.g. elevation and slope). For covariates only available before 2015 (e.g. poverty indices), we assumed that the general state or the geospatial relative ranking for that indicator did not change significantly over time. All covariate layers were prepared at a spatial resolution of 0.05 degrees (approximately 5km) due to the random displacement of DHS GPS cluster point locations (Burgert, Zachary, *et al*. 2013, Perez-Heydrich *et al*. 2013). Large water bodies identified in source data layers were excluded, retaining the same spatial extent for all covariate layers. To reduce collinearity, we excluded strongly correlated covariate pairs (|*r*| < 0.7), retaining the covariate in each pair least correlated overall with other covariates. Data pre-processing was performed using ArcGIS 10.4.1 (ESRI, Redlands, CA, USA).

### 2.3 Mapping prevalence of basic hygiene with an ensemble model

We adopted a model stacking (Wolpert 1992) approach to predict basic hygiene prevalence. Model stacking is a robust ensemble method that combines outputs of multiple modelling algorithms to improve prediction, in which the final combination rule is a generalised modelling algorithm instead of voting or averaging. The process of model stacking is often arbitrarily implemented via many levels, where the predictions generated by the modelling algorithms in a level become the inputs in the next level, until being generalised into the final prediction by the final level modelling algorithm (known as the ‘meta-model’ in machine learning). This ensemble approach has been applied in global mapping of basic hygiene at national level (Brauer *et al*. 2020) as well as demographic and health-related indicators of interest such as child growth failure (Osgood-Zimmerman *et al*. 2018), vaccine coverage (Mosser *et al*. 2019), and disease prevalence (Reiner *et al*. 2018, Dwyer-Lindgren *et al*. 2019). It has been shown to out-perform other conventional techniques (Clarke 2003). In this study, we adopted a multiple-level model stacking system as depicted in **Figure 2**. In the first level, we fitted five sub-models with the pre-processed cluster point data and geospatial covariate layers using five predictive modelling algorithms for regression problems: (a) Generalised Linear Model (GLM) (McCullagh and Nelder 1989); (b) Multivariate Adaptive Regression Splines (MARS) (Friedman 1991); (c) Support Vector Machines (SVM) (Vapnik 1995); (d) Classification and Regression Trees (CART) (Breiman *et al*. 1984); and (e) Gradient Boosting Machines (GBM) (Friedman 2001). These modelling algorithms were selected based on predictive accuracy, computational cost and ease of automatic parameter tuning in the R computational environment. The sub-model predictions generated using the geospatial covariates were then used as exploratory predictors in the next level meta-model for a generalised final prediction. We employed RF as the meta-model algorithm given its good predictive performance and advantages such as having fewer hyper-parameters to tune (Stevens *et al*. 2015). Since such machine learning predictive model does not account for spatial autocorrelation, the meta-model’s predictors additionally included a raster layer generated from sample cluster points using inverse distance weighting (IDW) interpolation (Philip and Watson 1982). This was based on the rationale behind the RFsp model (Hengl *et al*. 2018), but with buffer distances replaced by a simpler deterministic estimation method to reduce the computational cost. The outcome variable was therefore modelled as a function of the sub-model predictions and the inverse distance weighted interpolation. Model performance was evaluated by calculating the coefficient of determination (R-squared), root mean squared error (RMSE), and mean absolute error (MAE). In the case of poor meta-model accuracy (e.g. R-squared < 0.6), level 2 would be another round of sub-model fitting and prediction using the predictions generated by level 1 sub-models as inputs for noise-reducing (as highlighted in dashed lines in **Figure 2**), thereby making the meta-model level 3. The predictions generated by level 2 (repeated) sub-models were then used alongside the IDW-interpolated layer as inputs in the meta-model to produce the final generalised prediction with further improved accuracy.

**Figure 2.**
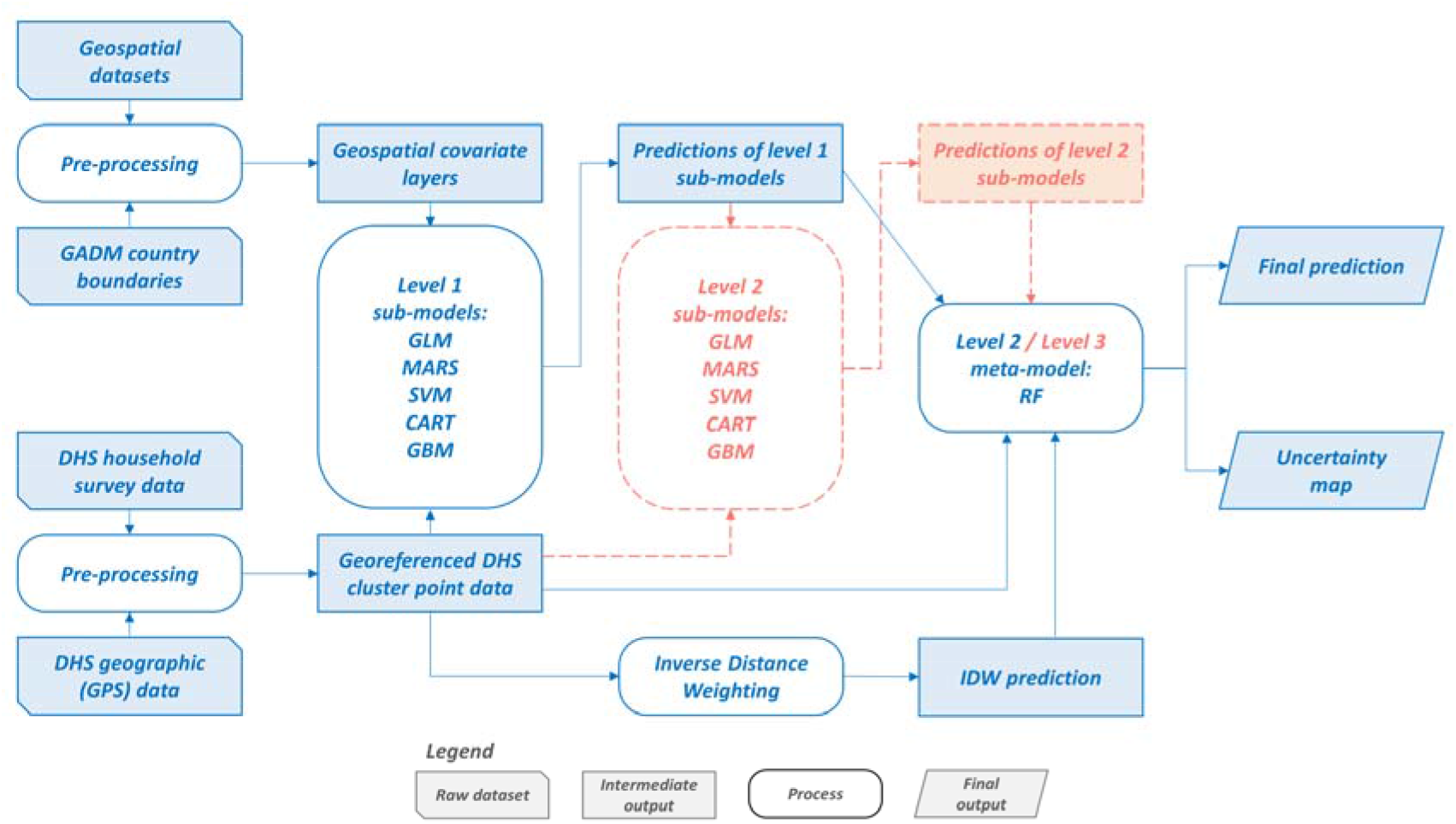
Flowchart of the model stacking methodology used in this study. Boxes in red dashed lines represent optional procedures adopted in the case of poor meta-model performance.

Model fitting, performance evaluation, prediction and analysis were carried out in R 3.5.2 terminal (The R Core Team 2020). The ‘caret’ R-package (Kuhn 2008) was used for automatic parameter sweeping, and performing Recursive Feature Elimination (RFE) for variable selection where no ‘built-in’ method was embedded in the sub-model algorithm’s R-package. RF models were fitted using the ‘randomForest’ (Liaw and Wiener 2002) package. All models were fitted using five-fold cross validation to avoid overfitting. Since our outcome variable was a proportion, an empirical logit transformation was performed before the meta-model. The logit transformation ensures that the final predictions can be converted back to a proportion value bounded by 0 and 1, so as to be easily interpreted and combined with population data for further analysis and validation.

As an additional means of model performance evaluation, we aggregated the generated prediction of each country to national and subnational boundaries depicted via publicly available data sources. Data on population, urban-rural divide and subnational level administrative boundaries were respectively derived from the WorldPop gridded population (Tatem 2017), the European Commission’s Global Human Settlement Layer (GHSL) Settlement Model grid (GHS-SMOD) datasets (Pesaresi *et al* 2019), the Global Administrative Areas (GADM) database v3.6 (Hijmans *et al* 2018), and the new Nepal administrative boundary database digitised by the Hermes GIS team fhttps://download.hermes.com.np/nepal-administrative-boundarv-wgs/: Accessed: 18^th^ June 2020). For countries where the corresponding DHS report (Uganda Bureau of Statistics (UBOS) and ICF 2018, National Institute of Population Studies (NIPS) [Pakistan] and ICF 2019, National Population Commission (NPC) [Nigeria] and ICF 2019) included estimates of basic hygiene coverage for subnational areas, Spearman’s Rho was employed to examine the correlation across subnational areal units between the DHS-reported coverage and our resultant estimates.

### 2.4 Examining spatial disparities in access to basic hygiene

To examine subnational disparities in basic hygiene services by area, the predicted prevalence of basic hygiene was aggregated to administrative level 2 for illustration by integrating with gridded population data (Tatem 2017). Using the resultant basic hygiene estimates, we calculated a dissimilarity index (Duncan and Duncan 1955, Yu *et al* 2014) as a measure of geographic inequality in access to basic hygiene services. This index measures the proportion of people in the total population who would have to shift location for basic hygiene services to be completely evenly distributed throughout all areas. The calculations were conducted at all available administrative levels, given the scale-dependency of this dissimilarity measure (Yu *et al* 2014). The administrative boundaries were derived from the GADM v3.6 database for most study countries, except Nepal where newly updated administrative boundaries digitised by the Hermes GIS team were used instead. Local contributions to national level inequality were also mapped, so as to reveal administrative level 2 areas contributing strongly to the overall disparity. Moreover, the predicted prevalence of basic hygiene was further broken down by type of human settlement following a classification system based on cluster population size, population density and built-up area density (Florczyk *et al*. 2019).

### 2.5 Ethics statement

Ethical clearance for this study was obtained from the Faculty of Social and Human Sciences, University of Southampton through the Ethics and Research Governance Online (ERGO) system (reference: 57472; approved on 17^th^ June 2020).

## 3. Results

For all study countries except Malawi, models were stacked in two levels. For Malawi, another round of sub-models was inserted in the middle of the total pipeline. Our ensemble models display good performance (all R^2^ above 0.8 except for Malawi where R^2^ = 0.61; see **Table S.3** in **Supplementaiy Material 1**), suggesting the majority of variance in the data was explained. The predictions at national and urban-rural levels are broadly in line with the figures in the DHS country reports (**Table S.4** in **Supplementary Material 1**), with significant increases in basic hygiene coverage found in both urban and rural Nepal. At subnational level, our estimates show patterns consistent with the DHS reported areal coverages (Nepal: r_s_=0.964, n=7; Nigeria: r_s_=0.891, n=37; Pakistan: r_s_=0.976, n=8; Uganda: r_s_=0.985, n=15; insufficient disaggregated data for Malawi). For the performance of sub-model algorithms, GBM is shown to out-perform the others in most cases according to the sub-model performance evaluation metrics (**Table S.3** in **Supplementary Material 1**). However, for most of the sub-model algorithms except GBM, this ranking is inconsistent with the contribution of sub-model predictions to the meta-model (**Table S.5** in **Supplementary Material 1**).

The relative contribution of the covariates varied by sub-model algorithm and by country (**Table S.6** in **Supplementary Material 1**). However, in most cases, a covariate with the highest importance in a sub-model also had high importance in other parallel sub-models. The only exception was improved water access, ranked first in the CART sub-model, but with moderate (ranked tenth out of 16 covariates) and low contributions (ranked number 14) respectively in the MARS and GBM sub-models. For Malawi, child growth failure (stunting) had the greatest contribution to the model. For Nepal, both night-time lights and women’s literacy were found to be the most influential covariates. Men’s literacy and access to improved water had the greatest contributions for Nigeria. In Pakistan, child stunting, women’s literacy and lack of sanitation (open defecation) were found most important In contrast, for Uganda, proportion of people living in poverty (defined by 2 USD per day) and child wasting had the greatest contributions. Across the five study countries and different sub-models, literacy (for men and/or women) provided useful information for modelling basic hygiene services in most cases (**Table S.6** in **Supplementary Material 1**). Surprisingly, in most cases, access to improved water did not provide useful information for modelling basic hygiene services. This is similar for lack of sanitation (open defecation) except in Pakistan.

**Figures 3-4** respectively show the spatially continuous estimates of the proportion of people in households with access to basic hygiene for each 5kmx5km grid cell for the five case study countries and corresponding uncertainty maps based on width between 95% confidence intervals. **Figure 5** shows the aggregated estimates for second administrative level areas, which highlights areas with high and low basic hygiene coverage. Household access to basic hygiene varies considerably by area - across geographic locations, by subnational areal units, and between urban-rural settings. Among the five case study countries, Malawi has comparatively little geographic variation in basic hygiene coverage, with all areas less than 50% at the 5km-grid level (**Figure 3A**), or less than 40% at the second administrative level (**Figure 5A**). Uganda, with a mostly rural population, also has poor basic hygiene services across most of the country, with relatively higher basic hygiene coverage in the south, particularly in areas bordering Lake Victoria. For the other three countries, basic hygiene coverage varies more geographically - for example, in Nepal, from 6.4% in Humla District, Karnali Pradesh to 93.1% in Lalitpur District, Bagmati Pradesh (**Figure 5B**). Pakistan has the highest coverage among the five study countries, with basic hygiene prevalent in northern Punjab, around Lahore (**Figure 5D**), and coastal areas, such as Karachi.

**Figure 3.**
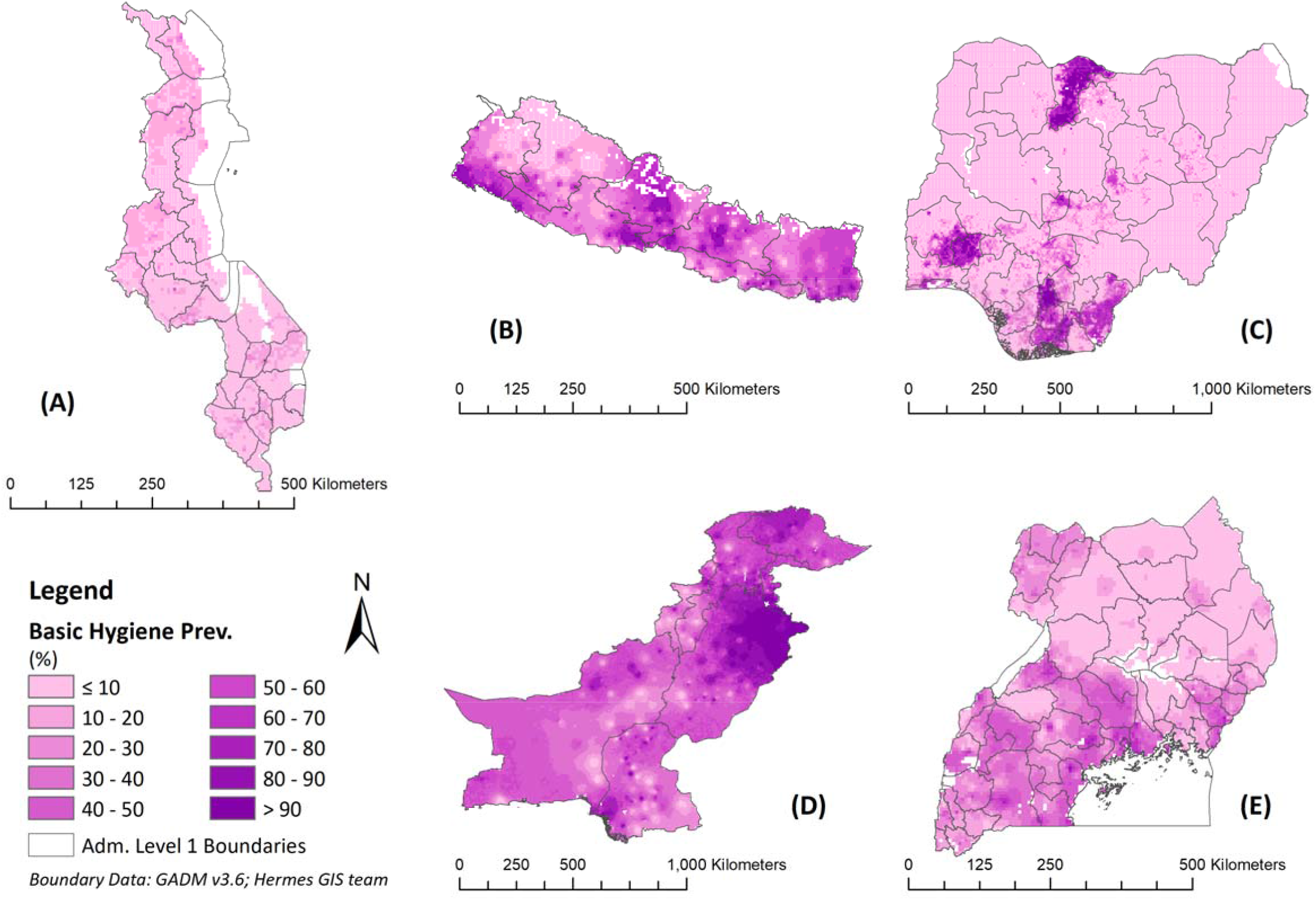
Modelled surfaces showing the estimated proportion of population living in a household with access to basic hygiene for (A) Malawi, (B) Nepal, (C) Nigeria, (D) Pakistan, and (E) Uganda

**Figure 4.**
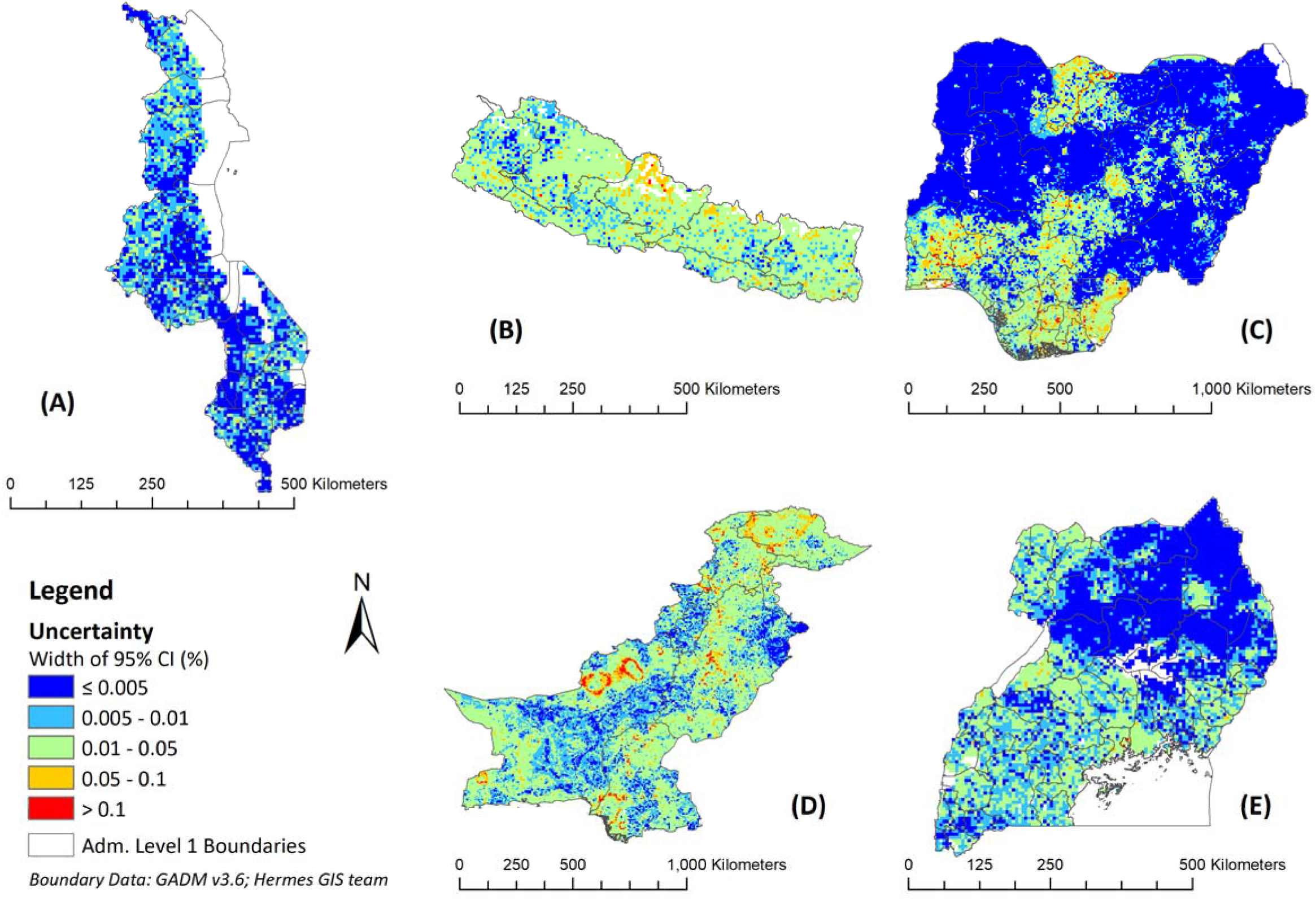
Uncertainty surfaces measured using the width of the 95% confidence intervals for (A) Malawi, (B) Nepal, (C) Nigeria, (D) Pakistan, and (E) Uganda

**Figure 5.**
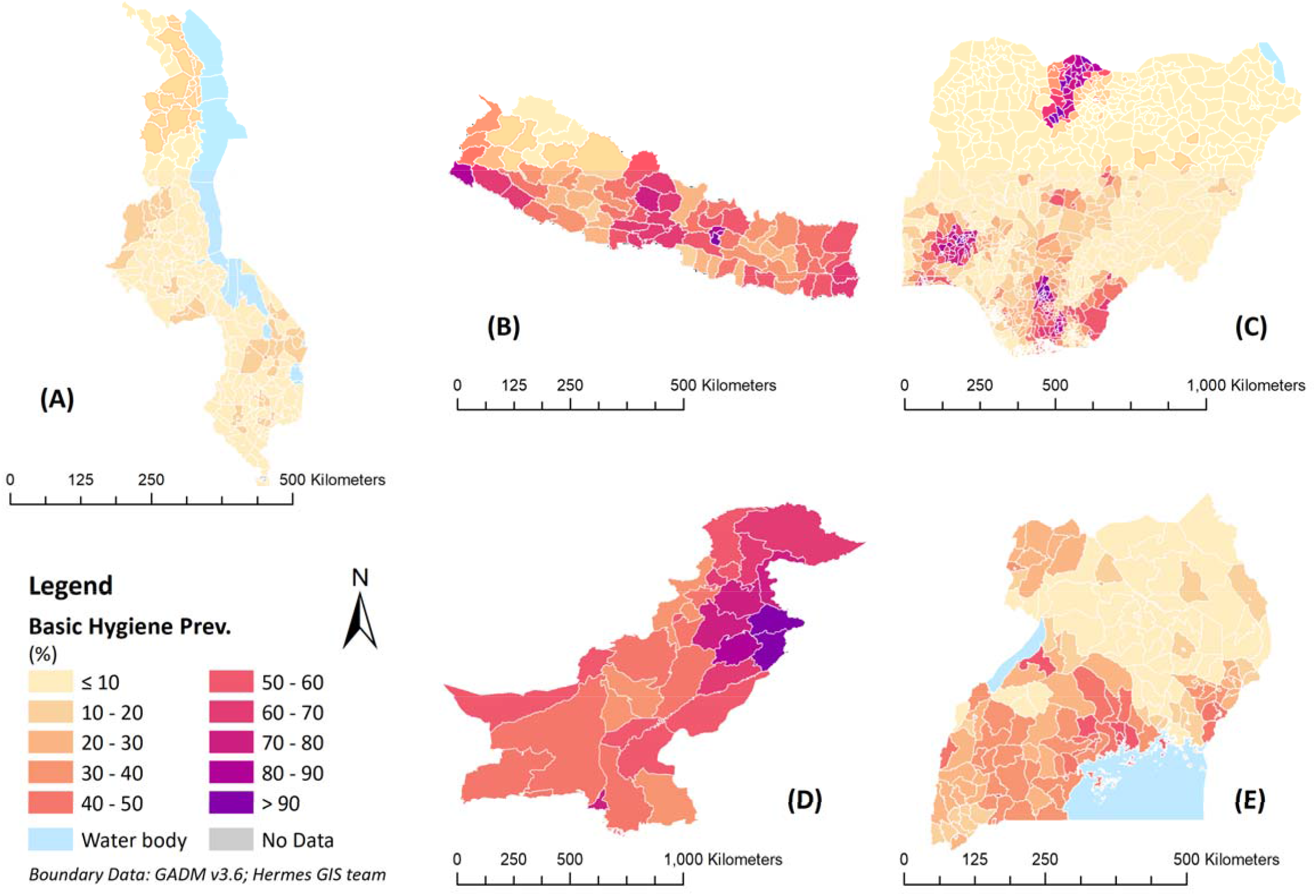
Estimated coverage of basic hygiene services at administrative level 2 for (A) Malawi, (B) Nepal, (C) Nigeria, (D) Pakistan, and (E) Uganda

Patterns in basic hygiene coverage (**Figure 5)** differed from the mapped local contributions to the national level inequalities (**Figure 6**). While the former measured the proportion of people with access to basic hygiene services, the latter measured the magnitude of segregation of population sub-groups with access versus those without access. For example, in Nigeria, the geographic disparities in basic hygiene services are pronounced with higher (> 60%) basic hygiene coverage concentrated in the south and Katsina State in the north, with very low (< 10%) coverage in most parts of northern and central Nigeria (**Figures 3C & 5C**). However, the map of local contributions to the dissimilarity index illustrated more spatially homogenous patterns, with stronger contributors also noticeable among areas where access to basic hygiene services was low, such as in the North East and North West. For the national level dissimilarity index, greater inequality in access is apparent in Nigeria (**Figure 7**), whilst Malawi, with the lowest levels of basic hygiene coverage, has consistently lower dissimilarity index values reflecting lower spatial inequalities in access relative to the other four countries.

**Figure 6.**
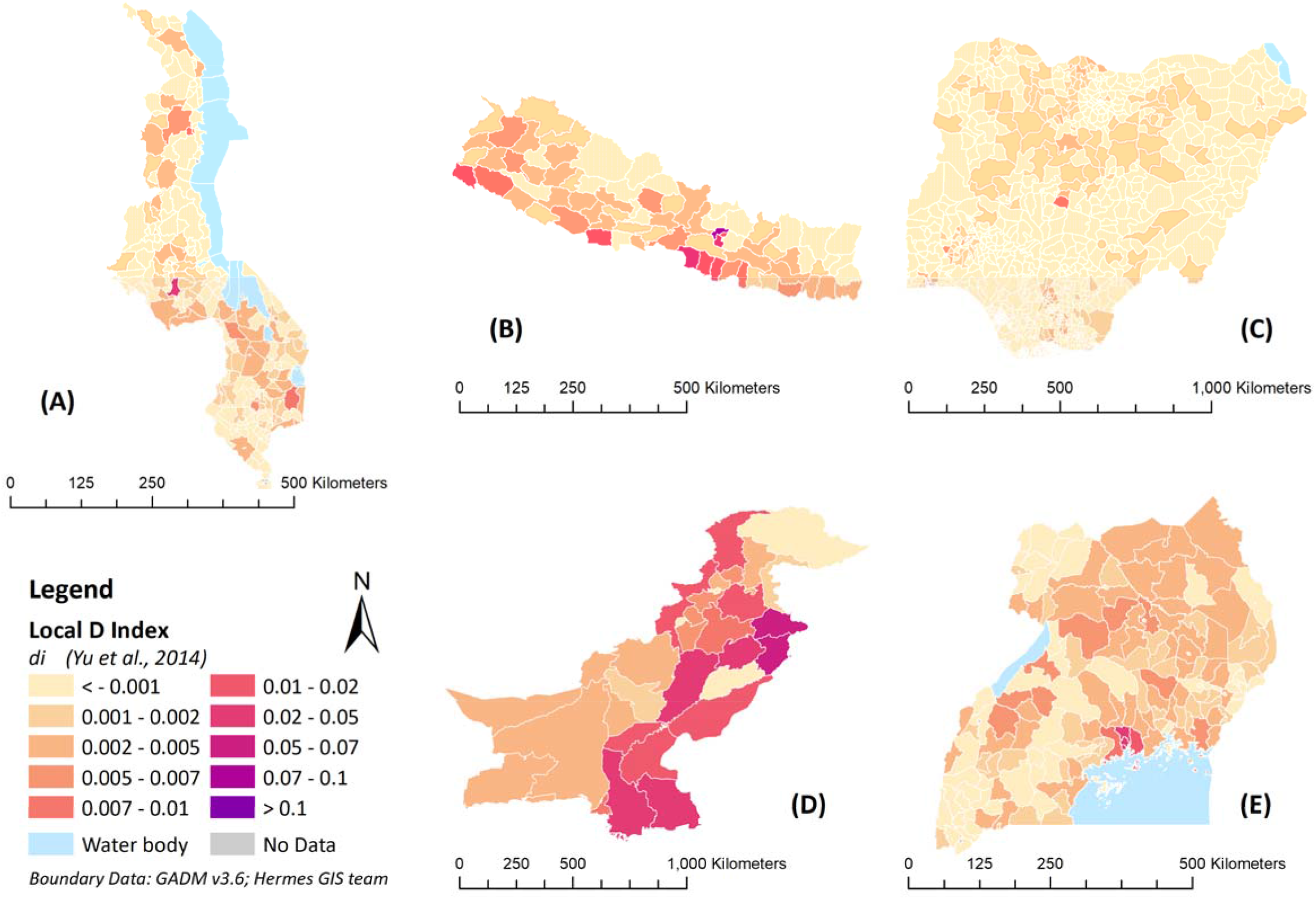
Map showing administrative level 2 local contributions to the national level dissimilarity index for basic hygiene services for (A) Malawi, (B) Nepal, (C) Nigeria, (D) Pakistan, and (E) Uganda

**Figure 7.**
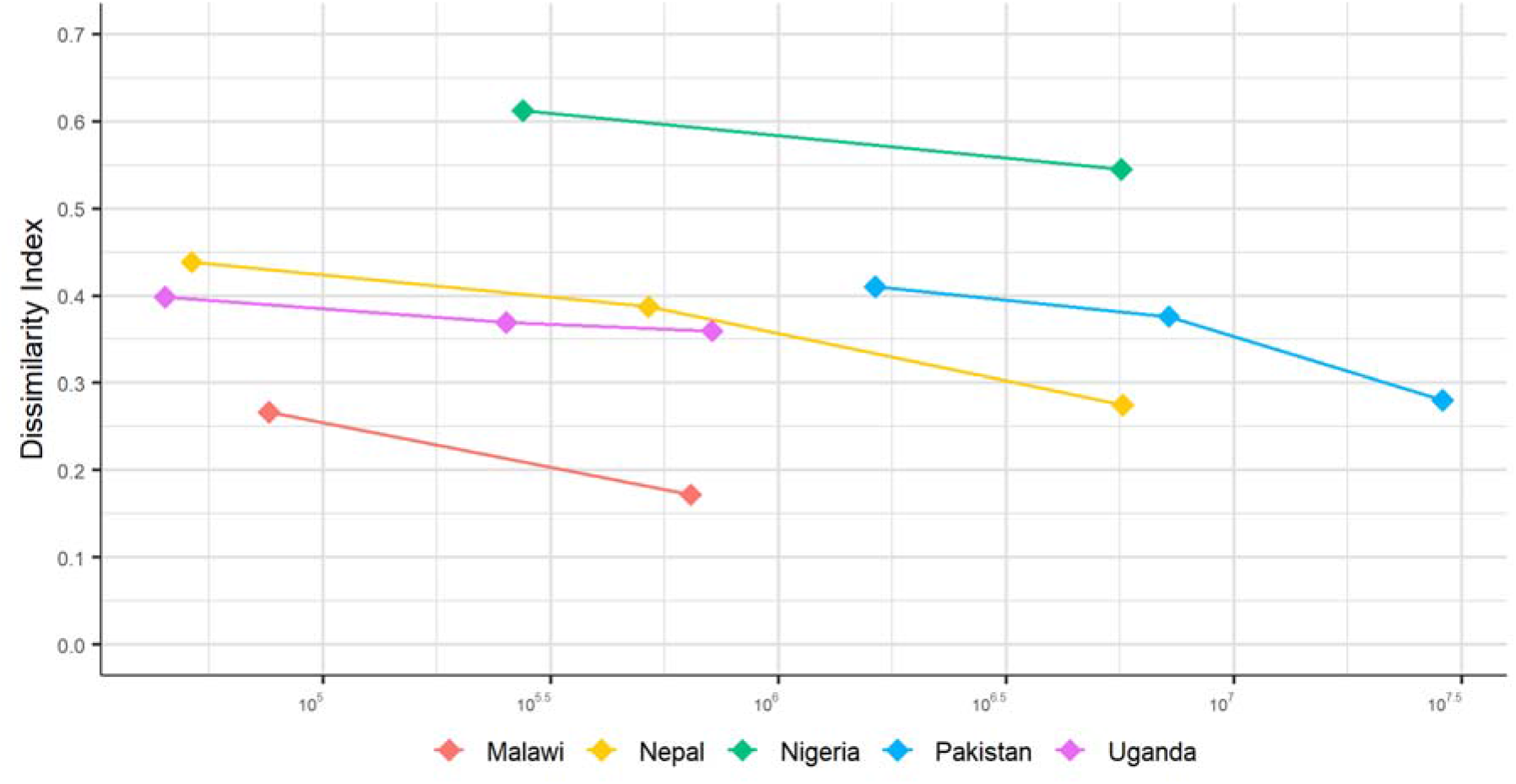
Dissimilarity indices for basic hygiene services Y-axis represents the value of the national level dissimilarity index for each administrative tier; X-axis is the mean population size of the administrative units (from administrative level 1 on the left to level 2 or level 3 on die right for each country) in base-10 log scale.

All five countries show gaps between urban and rural areas, with lower coverage apparent in rural areas in all cases. **Table 2** shows inequalities in access to basic hygiene services by type of human settlement. Pakistan has the highest coverage over all types of human settlement (mostly greater than 60%), with the lowest coverage in rural areas of very low density (56.9%). In many cases, the highest coverage of basic hygiene can be found in urban centres, except in Uganda (49.4% in suburb/peri-urban areas).

**Table 2.** Percentage of population living in a household with basic hygiene by type of human settlement

| Settlement Type | Malawi | Nepal | Nigeria | Pakistan | Uganda |
| --- | --- | --- | --- | --- | --- |
| Urban Centres | 14.2 | 78.9 | 52.1 | 78.7 | 47.5 |
| Dense Urban Clusters | 14.0 | 50.0 | 28.3 | 69.2 | 22.7 |
| Semi-Dense Urban Clusters | - | 42.9 | 21.0 | 64.6 | - |
| Suburb/Peri-Urban Areas | 6.2 | 44.0 | 49.9 | 71.7 | 49.4 |
| Rural Clusters | 7.7 | 44.3 | 24.1 | 69.5 | 30.4 |
| Low Density Rural Areas | 6.9 | 39.5 | 25.9 | 69.0 | 25.0 |
| Very Low Density Rural Areas | 6.7 | 51.6 | 17.4 | 56.9 | 21.5 |

## 4. Discussion

To our knowledge, this analysis represents the first nationwide estimation of basic hygiene prevalence in LMICs using machine learning predictive models, and represents a pioneering work examining geospatial disparities in access to basic hygiene services. In the context of the SDG targets for universal access to basic services for all by 2030 (WHO and UNICEF 2018), our results reveal substantial disparities in access to basic hygiene services across geographic locations. Notably, high levels of access to basic hygiene services often benefit people living in affluent areas, whereas the vast majority who lack access to basic hygiene live in economically disadvantaged communities. For example, in Nigeria, basic hygiene is more prevalent in coastal areas in the south, and less so in most of the north, home to 87% of the poor (World Bank 2020b). The substantial local disparities in coverage observed in this study suggest that estimates at national and provincial level are insufficient for monitoring progress towards universal access. While aggregated estimates often mask small pockets of low coverage, our modelled surfaces at the 5km-grid level and the adopted inequality metrics offer tools for a better understanding of the underlying local disparities in hygiene access hidden by national and regional averages.

Machine learning models such as RF have been adopted in various applications concerning spatial distributions. Such applications, either concerning environmental suitability or susceptibility across an area or looking for geographic weights for dasymetric population redistribution (Stevens *et al*. 2015), often ignore spatial autocorrelation (Hengl *et al*. 2018). Hence, studies looking at the prevalence of demographic and health-related indicators often draw on Bayesian geostatistical methodologies (Mayala *et al*. 2019). In this study, we employed a model stacking technique with several machine learning algorithms to predict basic hygiene prevalence, where an IDW interpolation was employed as an additional predictive covariate to account for spatial autocorrelation. The adoption of this IDW estimator incorporated geographical proximity effects into the model to ensure optimal prediction, and offered simplicity and a significant speed boost in computation. Both the performance evaluation metrics and the consistency between our predictions and DHS-reported figures suggest our resultant output is plausible. For Malawi, the comparatively little geographic variation in basic hygiene coverage with low prevalence in all areas may be the main reason for the relatively low model performance. The adoption of the model stacking technique was shown to improve model performance significantly, as reflected in the gain in performance evaluation metrics between levels (**Table S.3** in **Supplementary Material 1**).

As an additional benefit of the predictive machine learning technique, our ensemble model gained insights into landscape-level characteristics associated with basic hygiene prevalence. In this five country study, literacy had a high covariate contribution to basic hygiene prevalence in all countries except Uganda. This finding may suggest a link between educational attainment and demand for handwashing facilities and materials. Education has previously been shown to associate positively with water and soap presence at handwashing facilities (Loughnan *et al*. 2015) as well as handwashing behaviour (White *et al*. 2020). In this regard, reducing the educational attainment gap may help widen access to basic hygiene services in LMICs. In addition, household wealth is known to relate to handwashing behaviour and facility status in some domestic settings (Loughnan *et al*. 2015, White *et al*. 2020). This is directly and indirectly reflected in the high importance of covariates in modelling basic hygiene prevalence, including child growth failure for Malawi, Pakistan and Uganda; stable night-time lights for Nepal; and percentage of population living in poverty for Uganda. In contrast, despite the known association between water source availability and handwashing (White *et al*. 2020), access to an improved water surprisingly provided little useful information for modelling basic hygiene services in all countries except Nigeria. This may be caused by the inclusion of improved water sources that are non-piped, off premises or perceived as poor quality within this metric, since the proximity and perception of water sources also affect handwashing behaviour (Luby *et al*. 2009, White *et al*. 2020) and in turn may affect presence of handwashing facilities and materials. Similarly, while handwashing behaviour can often be linked to toileting and potential faecal contact (Wolf *et al*. 2019, White *et al*. 2020), lack of sanitation was only found among the most important covariates in Pakistan. This may also be because the sanitation indicator did not distinguish those improved sanitation types that have stronger associations with handwashing using water and soap (White *et al*. 2020).

Beyond local coverage gaps, a variety of inequality metrics have been employed in measuring geographic inequalities in water and sanitation access (Cetrulo *et al*. 2020), among which many can be decomposed at a regional level (Pullan *et al*. 2014, Yu *et al*. 2014, Chaudhuri and Roy 2017, He *et al*. 2018). Drawing on the decomposition of a dissimilarity index (Yu *et al*. 2014), this study examined the degree of segregation of the two groups of population with and without access to basic hygiene across geographic-sub-divisions. This dissimilarity index, through its calculation, offers simplicity and takes into consideration population size. It therefore provides additional information on the effect of population size hidden by local coverage. For example, in Malawi, a very high level of inequality in access to basic hygiene services in Lilongwe City can be found in the local dissimilarity index map (**Figure 6**), given its large population size as the capital city. In contrast, this contribution to overall inequality is masked in the local coverage map (**Figure 5**). This feature of the local dissimilarity index could provide a foundation for identifying population at risk, quantifying burden and guiding resource deployment by locating areas with high population density and low levels of access.

This study is subject to several limitations as follows: firstly, the precise coordinates for the DHS clusters were not available in order to protect respondents’ confidentiality (Burgert, Zachary, *et al*. 2013, Perez-Heydrich *et al*. 2013). The displacement of cluster locations restricts DHS spatial precision, and thus undermines the utility of the output in estimation at very fine spatial scales. Secondly, since handwashing has only recently been measured through household surveys, and since currently existing geospatial data sources on certain hygiene-related factors are limited, our study was based on a cross-sectional design using datasets as close to the present as possible. Temporal variation in basic hygiene unmeasured through these cross-sectional surveys may have reduced the strength of its association with the selected factors. Thirdly, our model may inherit the limitations and uncertainties of the input data and methods adopted in this study. This includes, for example, the inaccuracy caused by the 1% of further-displaced (up to 10km) rural cluster points in the model input; potential bias caused by the exclusion of samples in politically unstable areas from the DHS campaign (National Population Commission (NPC) [Nigeria] and ICF 2019); misleading information captured in stable night-time lights due to petroleum industry in certain areas such as the Niger Delta; potential inconsistency in urban-rural classification between our referenced data and the DHS-adopted definitions; potential effect of "no permission to see” in the DHS-reported figures; uncertainty in the geospatial data products used as predictive covariates; distortion caused by data pre-processing; and any drawbacks in the adopted modelling algorithms. In addition, the scale-dependency of the dissimilarity index undermines its utility in national comparisons, as the population size of subnational divisions varies between countries. Furthermore, this study was cross-sectional in design, which thus precludes causal inferences (Kesmodel 2018). Lastly, subject to context, the predicted prevalence of basic hygiene should be interpreted with caution, as the observed presence of basic hygiene items this study predicted would overestimate actual handwashing behaviour (Prüss-Ustün *et al*. 2019).

In this study, the machine learning algorithm RF was employed to generalise the final prediction in the stacked modelling framework. However, many existing efforts to map demographic and health-related indicators adopted a Bayesian geostatistical model using an ensemble approach, whereby the estimates reflect both geospatial and temporal dimensions. Subject to sufficient data being available, a future study could implement such an approach to produce estimates of basic hygiene coverage with extended temporal coverage. Such a study could systematically investigate the strengths and limitations of machine learning models in comparison with Bayesian geostatistical models. Furthermore, there would be scope to conduct a similar study examining predictors of basic hygiene services in other settings, including schools and healthcare facilities, which are priorities for SDG monitoring and infectious diseases prevention. In the context of the currently ongoing COVID-19 pandemic, there would also be scope to expand the analysis to other countries and to examine spatial patterns for hygiene in conjunction with other priority indicators.

## Conclusion

In view of geographical disparities in basic hygiene services, meeting the SDG target for universal access for improved public health requires monitoring at geospatially explicit scales. This study produced estimates of access to basic hygiene services at the 5km-grid scale for five low- and middle-income study countries using an ensemble model, reflecting the capability of machine learning and the value of existing geospatial datasets in predicting the prevalence of basic hygiene services. The methodology provided insights into geospatial patterns of basic hygiene services and their association with landscape-level characteristics. Both educational attainment and wealth status were found to be important in explaining the geospatial distribution of basic hygiene services. By triangulating with subnational administrative data, local coverage and inequality metrics were calculated to reveal apparent disparities in access to basic hygiene services, particularly highlighting areas with large populations. Such outputs can be used as alternative or supplementary information alongside the aggregated estimates. With extended geographic and temporal coverages in the future, they could become important tools to support planning of efficient and precise deployment to scale up access to hand hygiene facilities with water and soap and shift social and cultural norms on handwashing, and ultimately achieve universal access to basic hygiene and improved public health for all.

## Data Availability

All data used in the manuscript are publicly available and are listed in the supporting material.

